# AI Implementation in Safety Net Healthcare: Understanding Barriers and Strategies

**DOI:** 10.64898/2026.04.07.26350351

**Authors:** Ciera Thomas, Jee Young Kim, Alifia Hasan, Sena Kpodzro, Jennifer Cortes, Brianna Day, Shoshanna Jensen, Sean L’Huillier, Michelle O. Oden, Sara Zumbado Segura, Eric W. Maurer, Sonia Tucker, Sidney Robinson, Briseida Garcia, Elizabeth Muramalla, Sunny Lu, Neal Chawla, Manesh Patel, Suresh Balu, Mark Sendak

## Abstract

Safety net healthcare delivery organizations (SNOs) serve vulnerable populations but face persistent challenges in adopting new technologies, including AI. While systematic barriers to technology adoption in SNOs are well documented, little is known about how AI is implemented in these settings. This study explored real-world AI adoption in SNOs, focusing on identifying barriers encountered across the AI lifecycle and strategies used to overcome them. Five SNOs in the U.S. participated in a 12-month technical assistance program, the Practice Network, to implement AI tools of their choosing. Observed barriers and mitigation strategies were documented throughout program activities and, at the conclusion of the program, reviewed and refined with participants using a participatory research approach to ensure findings reflected lived experiences and organizational contexts. Key barriers emerged during the Integration and Lifecycle Management phases and included gaps in AI performance evaluation and impact assessments, communication with patients about AI use, foundational AI education, financial resources for purchasing and maintaining AI tools, and AI governance structures. Effective strategies for addressing these barriers were primarily supported through centralized expertise, structured guidance, and peer learning. These findings provide granular, actionable insights for SNO leaders, offering guidance for anticipating barriers and proactively planning mitigation strategies. By including SNO perspectives, the study also contributes to the broader health AI ecosystem and underscores the importance of participatory, collaborative approaches to support safe, effective, and ethical AI adoption in resource-constrained settings.

**Author Summary:** Safety net organizations (SNOs) are healthcare systems that primarily serve low-income and underinsured patients. While interest in artificial intelligence (AI) in healthcare has grown rapidly, little is known about how these organizations experience AI adoption in practice. In this study, we partnered with five SNOs over a 12-month program to document the challenges they encountered when implementing AI tools and the strategies they used to address them. We worked closely with SNO staff throughout the process to ensure our findings reflected their lived experiences with AI implementation. We found that the most common challenges arose when organizations tried to integrate AI into daily operations and monitor and maintain those tools over time. Specific barriers included difficulty evaluating whether AI was performing as expected, limited guidance on communicating with patients about AI use, a lack of resources for staff training, limited financial resources, and the absence of formal governance structures. Successful strategies for overcoming these challenges drew on shared knowledge and structured support provided by the program, as well as learning from peer organizations. These findings offer practical guidance for SNO leaders planning or managing AI adoption, and contribute to a broader conversation about what is required to implement AI safely and effectively in healthcare settings that serve the most medically and socially vulnerable patients.

## Introduction

Artificial Intelligence (AI) adoption in U.S. healthcare settings has accelerated rapidly in recent years. As many as 22% of U.S. healthcare organizations now use AI tools, which is more than double the 9% average adoption rate across all industries (1). Existing literature suggests that this growth is driven by mounting pressures on the industry, including clinician shortages and burnout, rising costs, operational inefficiencies, increasing public use of generative AI tools, and recognition of AI’s potential for supporting safer and more patient-centered care (1–3). However, adoption remains uneven across the healthcare delivery landscape, with significant variations across geographic regions, institutional size and type, ownership models, and resource availability (2,4,5). Larger organizations, those with systems affiliations, and those with strong digital interoperability infrastructure demonstrate higher implementation rates, and implementation experiences are well documented in these types of settings (4–9). In contrast, health delivery organizations (HDOs) in rural or medically underserved areas and those with provider shortages are significantly less likely to implement AI technologies (2,5). These disparities are especially visible among safety net healthcare delivery organizations (SNOs), where AI adoption rates remain comparatively low despite the significant potential benefits these tools could offer (10,11).

Such patterns of inequity in technology adoption reflect the growing digital divide, or the gap between individuals, households, businesses and geographic areas at different socio-economic levels with regard to their opportunities to access information and communication technologies (12). This divide can exacerbate disparities in health outcomes among the already vulnerable populations that SNOs serve, including rural communities, racial and ethnic minorities, and government-insured and self-pay patients (13–15). While AI adoption is increasing, little is known about its implementation in safety net settings. Understanding how AI is implemented in SNOs is therefore critical to mitigating the digital divide and ensuring that vulnerable communities can benefit from these technologies. The current study aims to explore real-world AI adoption in SNOs, with a focus on identifying barriers to implementation and strategies to address them.

### AI Adoption in Safety Net Health Delivery Organizations

SNOs such as Federally Qualified Health Centers (FQHCs), community hospitals, and community health clinics, are “providers who organize and deliver a significant level of health care and other needed services to uninsured, Medicaid and other vulnerable patients” (16). By offering healthcare services regardless of ability to pay, SNOs function as critical access points for over 34 million Americans who would otherwise lack regular access to care (17–19). SNOs serve a disproportionate percentage of low-income individuals, racial and ethnic minorities, uninsured or underinsured individuals, and rural communities compared to non-SNOs (19–21). In addition to clinical services, they also provide a variety of ‘wrap-around services’ such as interpretation and translation services, transportation, outreach, nutrition, and social support services which proactively seek to address social determinants of health (22). This comprehensive focus on serving vulnerable communities makes SNOs an important focal point for understanding and addressing health inequities, including in adoption of emerging health technology solutions.

Existing research suggests SNOs may stand to benefit most from adoption of health AI tools (10,11,13). Many SNOs experience acute staffing shortages while serving large volumes of patients with complex social and medical needs, creating conditions that are especially prone to provider burnout (15). Implementation of ambient AI scribes could help alleviate some of the documentation load and reduce this burden (11,23). Additionally, predictive analytics to identify chronic and acute conditions could mitigate disparities in early disease identification for underserved communities, improving health outcomes, reducing hospital admission rates, and streamlining referrals to specialty care (24,25).

Nevertheless, SNOs have demonstrated lower rates of AI adoption compared to other types of HDOs (4,5,7,11,13–15). These trends reflect persistent structural barriers, such as constrained funding for purchasing and maintaining AI tools, limited digital infrastructure and workforce capacity for supporting implementation, and insufficient access to legal and technical expertise needed for vendor contracting and risk assessment (11,13,15). Data limitations also pose a significant challenge. Fragmented and incomplete electronic health records (EHRs) make it difficult to train, validate, and implement reliable AI models, and existing multisite databases often fail to capture the demographic and clinical characteristics of safety net patient populations (13,15). This data gap creates additional concerns that the use of AI models trained on unrepresentative datasets could increase risk of algorithmic bias and perpetuate existing racial and ethnic disparities in health outcomes (11,14).

To support AI adoption in SNOs, early technical assistance efforts have begun to emerge. For example, the National Consortium of Telehealth Resource Centers—a network of 12 regional and 2 national centers dedicated to expanding access to care in rural and underserved communities through the implementation of telehealth programs—has recently expanded its scope to include supporting AI adoption (26). As part of this effort, one regional Telehealth Resource Center developed a Health Care AI Toolkit which is intended to provide guidance and resources for clinicians, operational staff, and provider leadership exploring AI solutions (27). For Fiscal Year 2025, HRSA included an elective objective for AI implementation in its Health Center Controlled Network (HCCN) funding program, thereby encouraging organizations to explore and expand the use of AI solutions (28).

These efforts represent important first steps for AI capacity-building efforts within SNOs. However, they primarily rely on one-time, resource-based supports rather than structured, longitudinal technical assistance models. As a result, they may be insufficient to address the multifaceted challenges associated with AI adoption in SNOs. This gap highlights the need for a more structured technical assistance model that integrates sustained support with context-specific guidance.

### Prior Technical Assistance Models

Slow uptake of AI within SNOs is not a novel pattern. At both the organizational and individual levels, SNOs have similarly experienced delayed adoption of other health technologies. A more structured approach to support technology adoption can be found in earlier initiatives that delivered technical assistance through coordinated networks, connecting centralized expertise to frontline organizations with sustained engagement.

One such example is observed with the adoption of EHRs. Prior to the implementation of the Health Information Technology for Economic and Clinical Health (HITECH) Act of 2009, EHR adoption lagged, with significant disparities observed in smaller, non-teaching, and rural hospitals (29–32). Given that EHR adoption yielded incremental but meaningful improvements in care processes, efficiency, and long-term value, disparities in EHR uptake risked perpetuating structural inequities by limiting under-resourced organizations’ ability to realize similar gains from emerging digital health infrastructure (33–35). To address the significant technical and organizational barriers that these HDOs faced, HITECH created and funded Regional Extension Centers (RECs) to provide technical assistance to these HDOs and accelerate EHR uptake (36–38). Provision of guidance on EHR product selection, privacy and security matters, and workflow redesign, among other services, resulted in 89% EHR adoption among REC participants compared to 58% among non-participants (39).

A similar technical assistance model was employed by Project ECHO, an initiative out of the University of New Mexico dedicated to building health system capacity and improving wellbeing globally (40). Project ECHO establishes Hubs, or specialist teams of experts across health, education, climate resilience, and system strengthening, to share expertise and engage directly with frontline workers (ECHO participants). Through these structures, frontline workers hone and acquire new skills through mentorship, collaborative problem-solving, and peer engagement under expert guidance. By continuing to cultivate and scale this community of practice, Project ECHO reached providers and educators in all 33 counties of New Mexico in 2025, equipping them with the knowledge, skills, and tools to serve their communities effectively and share best practices (41). Beyond New Mexico, the Project ECHO model has also been utilized to improve provider knowledge and capacity to treat multiple chronic conditions ranging from chronic pain to dementia in over 40 countries (42). Evaluations of initiatives using the ECHO model have shown demonstrable increases in provider knowledge and self-efficacy for treating chronic conditions as well as improved patient outcomes such as reduced 30-day readmission rates and emergency department visits (43,44).

RECs and the ECHO model both offer hands-on, tailored guidance rooted in ongoing mentorship, rather than one-off webinars or consultations. RECs offered site-specific guidance on selecting appropriate tools, adapting existing workflows, and ensuring compliance, among other services. ECHO Hubs cultivate ongoing mentorship and peer learning through routine virtual interactive community sessions and case-based training. These approaches highlight the importance of not just addressing the immediate technical challenge, but also providing ongoing education, building relationships and strengthening existing capacity to create sustainable improvements in knowledge and skills.

### The Practice Network Model for AI Technical Assistance

Health AI Partnership (HAIP) is a multi-stakeholder collaborative housed within the Duke Institute for Health Innovation that is composed of over 30 HDOs, ecosystem partners, and federal agencies. Its mission is to empower healthcare professionals to use AI effectively, safely, and ethically through community-informed up-to-date standards. Building on lessons learned from the success of the REC and Project ECHO technical assistance models, HAIP designed and implemented a 12-month technical assistance program, called the *Practice Network*, to help SNOs adopt AI tools of their choice using HAIP best practices.

The Practice Network Program was structured using a *distributed hub-and-spoke model* (37). Whereas traditional hub-and-spoke models like Project ECHO feature a single, central Hub providing technical assistance to Spokes seeking implementation support, the Practice Network employed a modified model with multiple Hubs to bring together interdisciplinary organizations with expertise across clinical, technical, operational, regulatory, and ethical domains. These Hubs provided health AI implementation guidance to Spokes—participating organizations without prior AI adoption experience. HAIP acted as both the *Coordinating Center*—organizing the program and coordinating activities across Hubs and Spoke sites—and as a Hub providing technical assistance.

In the 2024-25 Practice Network Program, 10 HAIP member organizations acted as Hubs with specialized AI expertise, including HDOs with prior AI implementation experience, an international law firm, an EHR vendor, a healthcare innovation nonprofit, and a community-based organization. The Spokes were five SNOs across the U.S. each pursuing first-time adoption of a specific AI tool within their organization. Hubs provided tailored technical assistance to each Spoke throughout the program, guiding them through implementation of their selected AI tools. To support this process, the Coordinating Center worked with Hubs to develop customized AI implementation plans for each Spoke, using the HAIP Key Decision Point framework, which maps best practices of AI adoption across four lifecycle phases: Procurement, Development/Adaptation, Integration, and Lifecycle Management (45). Each plan was tailored to the Spoke’s context, challenges, proposed AI tools, and adoption goals, serving as a roadmap for their activities throughout the program. Technical assistance was provided through regular progress check-ins, one-on-one mentorship, office hours, and peer networking events. As Spokes implemented their plans, they shared their learnings with both the Hubs and fellow Spokes, fostering a collaborative, practice-oriented learning environment. At the conclusion of the 12-month program (September 2024-September 2025), three Spokes had completed AI pilot testing and were conducting ongoing monitoring and evaluation of their tools. One Spoke launched pilot testing in October 2025, while the remaining Spoke completed a randomized controlled trial and transitioned its tool into full clinical operations in February 2026.

### Current Research

Research on AI adoption in SNOs remains sparse. Within this limited body of work, existing research has largely focused on structural or system-level challenges in technology adoption in SNOs, with limited attention to organization-specific, on-the-ground experiences of implementing particular AI tools. As a result, little is known about the practical barriers SNOs encounter during AI implementation or the strategies they use to address these challenges. To address this gap in the literature on AI implementation in safety net settings, this study examines real-world AI adoption within SNOs.

This study draws on a participatory research approach embedded within the Practice Network Program. As participants engaged in program activities, research staff documented observed barriers to AI implementation and emerging mitigation strategies. These initial findings were then reviewed, elaborated, and refined through iterative participant feedback to ensure that they accurately reflected participants’ lived experiences and organizational contexts. This process resulted in a set of empirically grounded barriers and corresponding strategies.

The primary objectives of this study are: (1) to understand the barriers to AI adoption that SNOs face during real-world implementation efforts across the AI lifecycle, and (2) to identify strategies employed to overcome these barriers. This study generates actionable insights that may inform other SNOs pursuing similar efforts. In addition, the findings contribute to the emerging literature on technical assistance for AI adoption in healthcare, offering evidence to inform the design of future programs that support safe, effective, and ethical AI implementation in safety net settings. By centering the perspectives of individuals leading AI adoption within SNOs, this study amplifies their voices and ensures they are represented within the health AI ecosystem.

## Methods

### Participants

A request for applications (RFA) was released in April 2024 to recruit SNOs across the United States for participation in the program. Organizations were considered eligible if they met several predefined criteria: (1) a stable EHR system, as the program did not support EHR implementation or migration; (2) secured leadership buy-in for AI adoption; (3) a designated implementation team consisting of a project manager, a clinical champion, and an IT champion; and (4) funding to procure an AI tool, as the program did not provide financial support for tool acquisition. The RFA remained open until August 2024, and applications were subsequently reviewed. As planned from the outset, the program selected five SNOs for participation in the inaugural cohort of the Practice Network program.

Selected SNOs (4 FQHCs and 1 community hospital) planned to adopt various types of AI tools, including two generative AI tools, two clinical decision support (CDS) tools, and one imaging tool. A total of 26 representatives from five organizations participated in the program, bringing expertise in technical (n=11), operational (n=11), clinical (n=14), and strategic (n=15) domains—where operational expertise refers to day-to-day process implementation and execution, and strategic expertise refers to organizational direction-setting and long-term decision-making. 22 participants had expertise spanning two or more areas. At the end of the program, 15 participants from the same organizations participated in focus groups as representatives of their organizations (Table 1). Detailed descriptions of each organization can be found in Appendix A.

**Table 1.** Overview of focus group participants across organizations.

| Organization | Location | Intended AI Use Case | Participants (n) | Participant Expertise Area |
| --- | --- | --- | --- | --- |
| Community-University Health Care Center | Minneapolis, MN | **CDS:** Predictive analytic for risk of patient no-show | 4 | Strategic*, Operational |
|  |  |  |  | Technical, Clinical |
|  |  |  |  | Technical, Clinical |
|  |  |  |  | Strategic, Technical |
| Health Center of Southeast Texas | Cleveland, TX | **Generative AI:** Ambient AI scribe | 2 | Operational, Technical |
|  |  |  |  | Technical |
| North Country Health Care | Flagstaff, AZ | **Generative AI:** Ambient AI scribe | 6 | Operational, Clinical |
|  |  |  |  | Operational, Clinical |
|  |  |  |  | Operational, Clinical |
|  |  |  |  | Operational, Clinical |
|  |  |  |  | Operational, Technical |
|  |  |  |  | Technical |
| San Ysidro Health | San Diego, CA | **Imaging:** Diabetic Retinopathy screening tool | 1 | Strategic*, Clinical |
| WakeMed | Raleigh, NC | **CDS:** Predictive analytic for risk of sepsis | 3 | Strategic*, Clinical, Technical |
|  |  |  |  | Strategic*, Technical |
|  |  |  |  | Operational |

\* Indicates executive-level individuals

### Procedures

A participatory research approach was employed to co-develop findings with participants and capture their real-world implementation experiences. The research process was structured into four sequential phases: (1) data collection, (2) inductive thematic analysis, (3) focus groups, and (4) content analysis.

#### I. Data collection

During the 12-month program, data were collected through field notes as participating sites progressed through stages of AI adoption. These notes documented activities completed, barriers encountered, and strategies utilized to overcome these challenges.

#### II. Inductive thematic analysis

An inductive approach was used to analyze field notes and identify barriers and strategies. One researcher reviewed and synthesized the notes, after which a thematic analysis was conducted to identify barriers encountered by participating organizations and corresponding mitigation strategies. Barriers across organizations were grouped based on conceptual similarity to identify recurring themes. Strategies utilized to address these barriers were identified through content analysis of field notes and listed with the corresponding barrier. Two types of strategies emerged during this analysis: (1) Spoke-initiated actions carried out independently by participating organizations, and (2) Hub-provided support through the Practice Network program, including mentorship and peer learning.

#### III. Focus groups

Structured focus groups were conducted with representatives from each participating organization to validate the findings of thematic analysis. Prior to each focus group, customized, interactive slide decks were created for each organization, following a standardized format (Fig 1). These decks outlined barriers encountered by each organization and strategies utilized to address them across different stages of AI adoption. They also allowed participants to provide real-time feedback.

**Fig 1.**
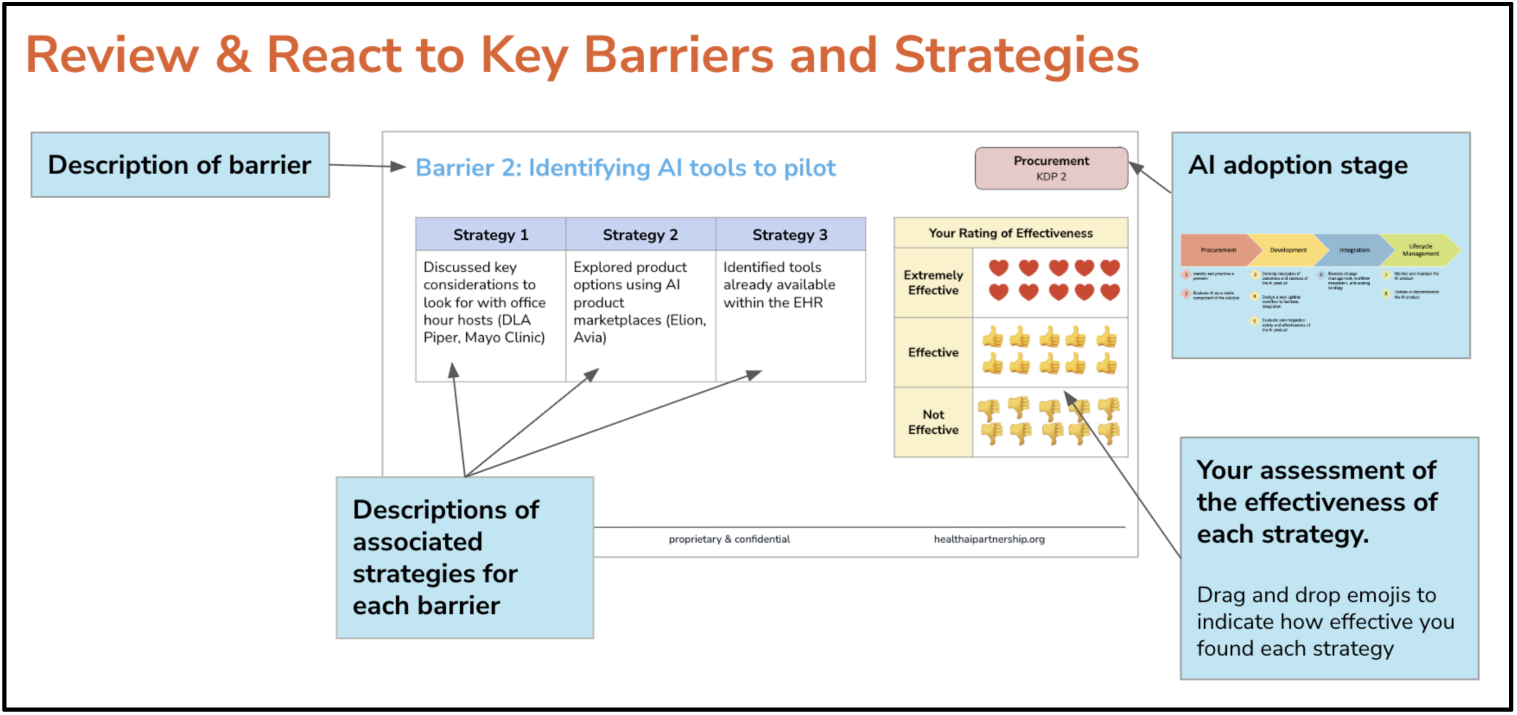
Format of focus group slides used to present barriers and associated strategies for participant validation.

Each focus group was conducted virtually, lasted approximately one hour, and was facilitated by two researchers. During the session, participants were presented with the slide deck and first asked to confirm whether each barrier resonated with their AI adoption experience. Subsequently, they were asked to review each strategy and rate its effectiveness using one of three emojis: a *thumbs-up* to indicate the strategy was effective in addressing the associated barrier, a *thumbs-down* to indicate it was not effective, and a *heart* to indicate the strategy was extremely effective. For each emoji selection, participants were prompted to explain their reasoning and describe any contextual factors (i.e., resource constraints, technical capabilities, end user buy-in) that may have affected the effectiveness of the strategies. If a presented strategy was not applicable, participants were asked to verbally acknowledge it and not select an emoji. Focus group sessions were recorded, and researchers took detailed notes to supplement the recordings for analysis.

#### IV. Content analysis

Content analysis was conducted to identify the most common barriers and evaluate the effectiveness of mitigation strategies. This approach provided a systematic way to synthesize the insights gathered during the focus group sessions. Two primary metrics were developed to guide this analysis: *barrier frequency* and *strategy effectiveness*.

*Barrier frequency scores* measure the number of participating organizations that encountered a specific barrier and represent how commonly each barrier was experienced across participating organizations during the program. Thus, scores range from 1 to 5, with higher values indicating greater prevalence. A score of 1 reflects a barrier unique to a single organization or its specific use case, whereas a score of 5 indicates that all five organizations encountered the barrier. Given the diversity of implementation contexts—including variations in AI application type (e.g., large language models vs. clinical decision support tools), organizational structure (single-site vs. multi-site health systems), and geographic location—barrier frequency reflects contextual specificity rather than sample size. Lower frequency barriers are not less reliable findings; rather, they represent context-specific challenges relevant to particular implementation scenarios.

*Strategy effectiveness* represents the perceived effectiveness of each strategy and were derived from participants’ emoji ratings. Strategies that were perceived as not effective were given a thumbs down and assigned −1 point, those that were perceived as effective were given a thumbs-up and assigned +1 point, and those that were considered very effective were given a heart and assigned +2 points. Strategies that were considered but not ultimately used were given a score of 0 as they were perceived to be neither effective nor ineffective. Strategy effectiveness scores were calculated in two steps. First, organization-level scores were computed by dividing the total points assigned to each strategy by the number of respondents per organization. This approach gave equal weight to each organization regardless of team size, preventing larger teams from disproportionately influencing results and maintaining fidelity to the program structure in which each organization represented a distinct implementation context. Second, these scores were aggregated across organizations by calculating the mean of organization-level scores—the sum of organization-level scores divided by the number of organizations that encountered the barrier (equal to the barrier frequency score). For example, if three organizations encountered a barrier and their organization-level scores for Strategy A were 1.5, 1.0, and 2.0, the strategy effectiveness score would be (1.5 + 1.0 + 2.0) / 3 = 1.5. Since strategy effectiveness scores are based on participants’ subjective ratings, they represent a continuous gradient of perceived strategy effectiveness rather than a binary measure. Scores range from −1 to 2, where values below 0 indicate a strategy was *not effective* in overcoming the barrier, values from 0 to <1 indicate a strategy was *minimally effective* in overcoming the barrier, values from 1 to <2 indicate a strategy was *effective* in overcoming the barrier, and a value of 2 indicates a strategy was *very effective* in overcoming the barrier.

### Ethics Statement

The present research was considered a quality improvement project and did not constitute human subjects research. Therefore, it was exempt from IRB review and approval at Duke University Health System. All participants provided verbal consent to participate in the research and to allow anonymized data to be used in analyses.

## Results

### Overview of Findings

A total of 21 barriers to AI implementation were identified across the lifecycle, with the majority occurring during the Integration phase and the smallest number occurring during the Development/Adaptation and Lifecycle Management phases (Table 2). Specifically, five barriers were documented during Procurement (two unique barriers with a barrier frequency score of 1 and three common barriers with a barrier frequency score of 3 or greater), four during Development/Adaptation (three unique barriers with a barrier frequency score of 1 and one barrier with a barrier frequency score of 2), eight during Integration (one unique barrier with a barrier frequency score of 1 and five common barriers, four of which had a barrier frequency score of 3 or greater), and four during Lifecycle Management (one unique barrier with a barrier frequency score of 1 and three common barriers, two of which had a barrier frequency score of 4 or greater). Barriers in the Procurement phase reflected difficulties in understanding the problem to be addressed by the AI tool, identifying product options, and interacting with vendors to procure tools. In the Development/Adaptation phase, barriers were largely associated with silent validation procedures and performance, as well as adapting workflows for AI integration. Barriers in the Integration phase included educating end users, change management challenges, communicating AI-related information to end users and patients, establishing processing for monitoring and evaluation, and model performance issues. Finally, barriers in the Lifecycle Management phase represented challenges with building internal capacity for maintaining and scaling AI tool implementation.

**Table 2.**
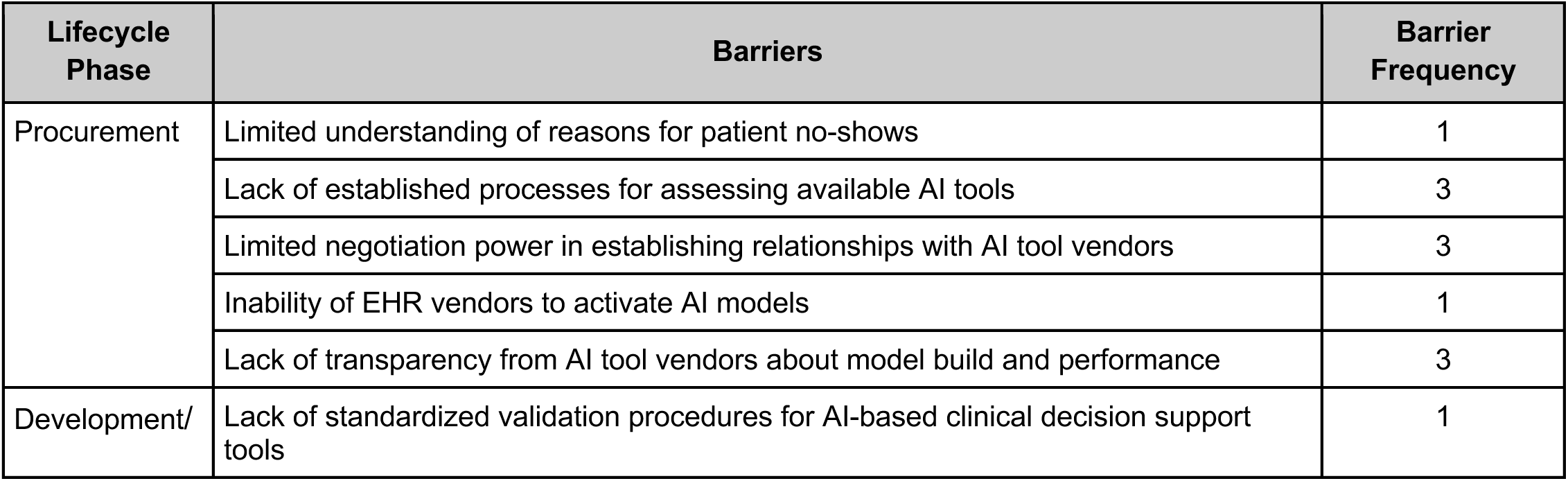
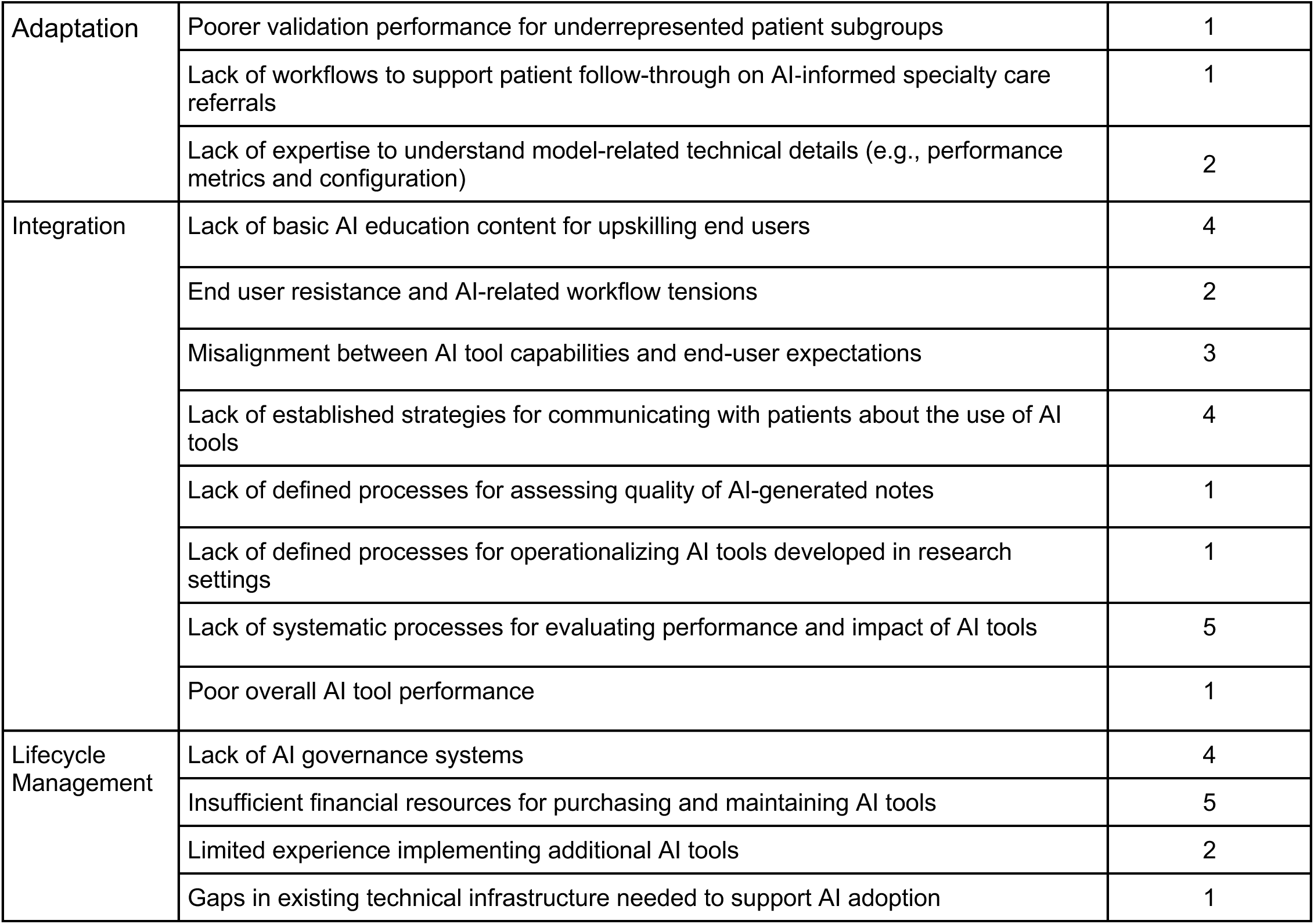
Barriers identified across the four phases of the AI lifecycle.

In total, 58 strategies to address these barriers were observed across the four lifecycle phases (Table 3; see Appendix B for a full list of strategies associated with each barrier). Of these strategies, 31 were Spoke-led actions carried out independently by participating organizations, and 27 were Hub-provided support through the Practice Network program. Each barrier was associated with 1 to 8 strategies, with a median of 2. Most strategies were identified in the Integration phase (n = 27), followed by Procurement (n = 13), Lifecycle Management (n = 12), and Development (n = 6). Perceived effectiveness of these strategies varied across lifecycle phases (Table 3). Participants rated strategies in the Lifecycle Management (mean = 1.0) phase as effective, while those in Procurement (mean = 0.7), Development/Adaptation (mean = 0.8) and Integration (mean = 0.9) were rated as minimally effective. Across all phases, strategies were largely rated as effective or minimally effective. The only exception was one strategy in the Procurement phase, developed by a Spoke, which addressed a barrier unique to that Spoke and was rated not effective (strategy effectiveness score = −1).

**Table 3.** Overview of barriers and strategies identified across four phases of the AI lifecycle.

| Lifecycle Phase | Number of Barriers per Phase | Number of Strategies per Phase | Strategy Effectiveness Mean | Strategy Effectiveness Median | Strategy Effectiveness Min | Strategy Effectiveness Max |
| --- | --- | --- | --- | --- | --- | --- |
| Procurement | 5 | 13 | 0.7 | 1 | -1 | 1.7 |
| Development/Adaptation | 4 | 6 | 0.8 | 1 | 0 | 1 |
| Integration | 8 | 27 | 0.9 | 0.8 | 0 | 2 |
| Lifecycle Management | 4 | 12 | 1.0 | 1 | 0 | 2 |
| *Total* | *21* | *58* | *--* | *--* | *--* | *--* |

### Most Common Barriers and Associated Strategies

There were five barriers which received a barrier frequency score of four or higher, signifying that they were experienced by the majority of Spokes (Table 2). These barriers were observed in the Integration and Lifecycle Management phases. To address these barriers, 18 strategies—almost a third of all strategies identified through this research—were used. Among these strategies, seven represented actions carried out within the Spokes, while the remaining 11 reflected support provided by the Hubs. One of these strategies was rated very effective, eight were rated effective, and nine were rated minimally effective. None were rated as not effective. Table 4 presents the most common barriers and associated strategies, which are described in greater detail below.

**Table 4.** Most common barriers experienced by Spokes and associated strategies, sorted by strategy effectiveness score.

| Lifecycle Phase | Barrier and Frequency (n) | Effective Strategies |  | Minimally Effective Strategies |  |
| --- | --- | --- | --- | --- | --- |
|  |  | Strategy Description | Strategy Effectiveness Score <sup>1</sup> | Strategy Description | Strategy Effectiveness Score <sup>1</sup> |
| Integration | Lack of basic AI education content for upskilling end users (n=4) | Accessing educational events, materials, and online courses shared in the Practice Network Program Resource Library | 1.4 | Employing training materials provided by the AI tool vendor | 0.6 |
| Integration | Lack of established strategies for communicating with patients about the use of AI tools (n=4) | Discussing disclosure versus consent considerations and applicable regulations with Hubs | 1.4 | Peer-learning through collaborative exchange of communication materials and strategies between Spokes | 0.8 |
|  |  |  |  | Adapting disclosure script template provided in the Practice Network Program Resource Library | 0.7 |
|  |  |  |  | Reviewing patient surveys and a disclosure script drafts created by the organizations with Hubs | 0.7 |
| Integration | Lack of systematic processes for evaluating performance and impact of AI tools (n = 5) | Learning from other participating Spokes by exchanging performance metrics and project plans | 1.4 | Partnering with academic medical centers or AI tool vendors to support performance evaluations | 0.6 |
|  |  | Discussing the results of performance evaluations conducted by the AI tool vendor with Hubs | 1.4 |  |  |
|  |  | Discussing pilot and post-deployment monitoring approaches and strategies with Hubs | 1.0 |  |  |
| Lifecycle Management | Insufficient financial resources for purchasing and maintaining AI tools (n=5) | Setting clear performance metrics for demonstrating return on investment (ROI) | 1.0 | Identifying grant funding opportunities | 0.4 |
|  |  |  |  | Negotiating pricing plan structure with AI tool vendors | 0.2 |
| Lifecycle Management | Lack of AI governance systems (n=4) | Utilizing data governance resources in the Practice Network Program Resource Library | 2.0 | Developing AI tool evaluation rubrics | 0.9 |
|  |  | Strategy Description | Strategy Effectiveness Score <sup>1</sup> | Strategy Description | Strategy Effectiveness Score <sup>1</sup> |
|  |  | Targeted guidance from Hubs on developing and operationalizing AI governance strategies | 1.8 | Polling internal staff and leadership to identify AI-related attitudes, priorities, and concerns | 0.5 |
|  |  | Reviewing AI adoption and use policy drafts with Hubs | 1.5 |  |  |

<sup>1</sup> Strategy Effectiveness scores range from -1 to 2, with <0 being *not effective*, 0 to <1 being *minimally effective*, 1 to <2 being *effective*, and 2 being *very effective* in overcoming the barrier.

#### I. Lack of basic AI education content for upskilling end users

While preparing end users for the adoption of AI tools in the Development/Adaptation phase, four Spokes expressed frustration with the limited availability of educational content on foundational AI concepts designed for frontline healthcare professionals. Participants reported that much of the publicly available content was either too advanced or too time-intensive to be practical, while paid content was often cost-prohibitive. Content provided by AI tool vendors was considered a minimally effective strategy. Participants noted that while vendors often supply basic training materials for their specific tools, these materials often fail to cover the foundational AI knowledge that end users need prior to implementation.

Curated resources in the Program Resource Library, including publicly available educational courses, handouts, and webinars, proved an effective strategy for addressing this gap. These materials and events were valued for being succinct, freely accessible, and focused on discrete, practical topics (e.g., governance, vendor evaluation, and monitoring), often accompanied by brief reference handouts that participants could revisit asynchronously. Nevertheless, participants expressed a continued desire for more streamlined, introductory educational content than currently available. Thus, this barrier was not fully resolved. As one participant shared, “I wish we actually got some more concrete guides for basic education. [… ] That’s something that I think could be a little bit better.”

#### II. Lack of established strategies for communicating with patients about the use of AI tools

In the Development/Adaptation phase, four Spokes reported difficulty determining how to communicate the use of AI tools to patients. Spokes faced two main challenges with regard to patient communication: deciding whether to seek disclosure or informed consent, and developing standardized materials to adequately inform patients about the use of AI tools in their care and assess their attitudes toward its use. All Spokes that encountered challenges in communicating the use of AI tools to patients ultimately addressed this barrier by developing processes for communicating with patients about the use of AI tools in their care.

To guide decision-making about seeking disclosure or consent, Spokes reported that discussing relevant considerations and applicable regulations with the Hubs was an effective strategy. Hubs provided guidance on key factors to consider when choosing whether to disclose AI tools use or seek informed consent, including state-level regulations, data sensitivity, impacts on patient privacy and trust, and operational implications. Ultimately, all Spokes chose to disclose AI tool use and provide patients with the opportunity to opt-out instead of seeking informed consent. Participants described choosing disclosure over informed consent because obtaining meaningful consent would require providing a level of AI education that was not feasible within clinical workflows. At the same time, disclosure was seen as necessary to uphold patient trust. This consideration carries particular structural weight in FQHCs, as federal mandate requires that 51% of board seats be held by patients, the remainder are held by community members (46).

Once decisions to disclose AI use had been made, Spokes faced the challenge of how to communicate this disclosure. They employed three primary strategies to address this challenge: adapting a generic disclosure language template, engaging in peer learning, and seeking Hub review of communication materials. To support these efforts, Spokes developed a set of patient-facing communication materials tailored to different communication needs. These materials included provider disclosure scripts, which were designed to give clinicians standardized language for disclosing the use of AI tools during patient encounters; informational handouts, which were intended to provide patients with additional detail about the AI tools being used in their care; and patient surveys, which were developed in recognition of the central role patients play in advising SNO operations and the priority SNOs place on maintaining patient trust and understanding patient attitudes toward AI use. The surveys were used to assess patient satisfaction before and after AI implementation and to collect feedback about the tools. In developing these materials, some Spokes drew directly on a generic disclosure language template made available by the Coordinating Center through the Program Resource Library when drafting their disclosure scripts. Spokes also benefited from informal exchanges with peer Spokes working on similar use cases, sharing and comparing disclosure scripts and patient surveys as a way to refine their approaches. Draft communication materials, including scripts, handouts, and surveys, were also shared with Hubs for review and additional feedback.

Spokes reported that strategies for communicating disclosure were minimally effective. This was largely because Spokes were working on different AI use cases and were at different stages of implementation. The diversity of clinical contexts and AI applications limited the extent to which disclosure scripts, handouts, or surveys could be easily shared or adapted across sites. In addition, some Spokes had not yet reached the point in implementation where patient-facing disclosure was required. While some Spokes were actively considering how AI use might eventually be communicated to patients (i.e., disclosure or informed consent, communication tools that might be used), they had not progressed to drafting materials, which reduced opportunities for meaningful peer learning or feedback. As a result, the practical impact of shared templates, peer exchange, and Hub review was more constrained.

#### III. Lack of systematic processes for evaluating performance and impact of AI tools

In the Development/Adaptation phase, all participating Spokes faced difficulties with systematically evaluating the performance and impact of AI tools, both during the pilot phase and following integration. To overcome this barrier, participants engaged in Hub mentorship and peer learning. For Spokes that lacked staff with AI expertise, Hubs assisted in interpreting results of performance evaluations conducted by the AI tool vendor to ensure the tool performed safely and effectively in the Spokes’ setting. Additionally, for ongoing post-integration monitoring, Hubs provided guidance on identifying appropriate metrics to measure AI tool success and on developing systems to collect both baseline and post-implementation data on these metrics. Among Spokes with the same use case, participants exchanged metrics and evaluation plans. While these strategies were reported as effective, challenges with implementing these systematic processes remained. At some Spokes with less technical staff, participants indicated that the barrier still persisted at the conclusion of the program, noting that implementing these assessment processes required more human resources than were available. As one participant stated, “I discussed the evaluation approach with our site mentor, talking about the metrics that we can use within the EHR. I think that was effective because the ideas and strategies were great. But we’re still struggling with having the extra staff to help us run these reports and create these things.”

Another proposed strategy was formalizing partnerships with regional AMCs or AI tool vendors to support evaluation of more complex use cases. However, this approach proved challenging because of insufficient local partnerships, limited access to data and tools, and a broader lack of AI-related knowledge and skills within the regional care delivery environment. One participant said, “We need to figure out locally how to continue developing these relationships or get support to continue to move forward… For some of us that don’t already have an academic affiliation or similar sites nearby, these data pieces and the use of data-supported clinical decision tools are really hard to imagine without having a more formalized relationship with local or regional partners.” This strategy was therefore considered minimally effective rather than effective.

#### IV. Insufficient financial resources for purchasing and maintaining AI tools

The financial resources required to purchase and maintain AI tools represented a significant barrier for all Spokes that was not fully overcome. As most Spokes operated with narrow margins, this financial pressure meant that these Spokes needed AI tools to quickly demonstrate fiscal return on investment (ROI) to justify the associated costs. As one participant described, “It’s just so hard, because what our team wants to see is this immediate ROI. But I really do think we’ve shown that there is expected long-term benefit with recruitment and retention.” Although participants assessed soft ROI in the form of non-fiscal impacts of AI tool use, formal metrics to evaluate and demonstrate financial ROI were not collected. Participants therefore identified the development of clear financial ROI metrics as an essential strategy for future implementation efforts and labeled it an effective strategy.

To enable initial investments and sustain fiscal requirements beyond the pilot period, some Spokes sought relevant grant opportunities or attempted to negotiate pricing arrangements with AI tool vendors to reduce cost, contract durations, or minimum user requirements. These strategies proved to be minimally effective, as organizations struggled to identify funding opportunities and vendors were often reluctant to adjust pricing structures.

#### V. Lack of AI governance systems

In the Lifecycle Management phase, four Spokes identified the absence of internal AI governance structures as a critical barrier to AI adoption. All Spokes that encountered this barrier successfully established governance policies and systems through resources from the Program Resource Library and mentorship from the Hubs. One participant reflected, “From my perspective, this was the biggest success of our participation in the program—all the work we were able to get done with governance. I honestly sometimes can’t believe how much progress we made in getting things set up and getting them board-approved.”

Various strategies were employed to help Spokes build AI governance structures. Because many Spokes lacked the foundational data governance processes required to build AI governance systems, resources on this topic were curated in the Program Resource Library. Participants found these resources particularly useful, making this the only strategy associated with the most common barriers to be rated as very effective. Other effective strategies for building internal AI governance systems included feedback from the Hubs on AI adoption and use policies drafted by Spokes, as well as guidance from the Hubs on operationalizing these policies. The Hubs advised Spokes on creating product request intake forms and governance committees and provided suggestions on who should serve on these committees, what their roles and responsibilities should be, and what their scope of work should include.

One minimally effective strategy focused on creating AI product evaluation rubrics to systematically assess available tools as part of AI governance development. The HAIP AI Vendor Disclosure Framework and other evaluation rubrics used by Hubs were provided in the Program Resource Library to inform this effort (Kpodzro et al., 2025). Using these resources, Spokes developed their own rubrics for assessing vendors on key issues such as secondary data use and data storage and security. However, challenges remained. Spokes reported that vendors often refused or were reluctant to complete these assessments, which prevented them from conducting a proper evaluation of the available tools. When vendors did respond, the information provided was often either too superficial to be useful or too technical for staff without AI expertise to interpret, further underscoring the need for AI education.

To inform AI policies and processes, another minimally effective strategy involved surveying internal staff and leadership to assess their attitudes, priorities, and concerns about using AI tools. The effectiveness of this strategy depended on when surveys were administered during the AI adoption journey. One Spoke distributed its survey prior to implementation and found the responses helpful for informing AI policy development. In contrast, other Spokes conducted these surveys after implementing AI tools and rated the responses not effective, describing persistent difficulties with securing leadership engagement and building end-user buy-in.

## Discussion

This study identified barriers and associated mitigation strategies that SNOs experienced across the AI lifecycle through real-world implementation initiatives. The most common barriers emerged primarily during the Integration and Lifecycle Management phases. Across these barriers, shared challenges emerged in two broad areas: (1) organizational capacity and resource constraints, and (2) the development of best practices for implementing and overseeing AI use. Regarding capacity and resources, participants emphasized the need for financial resources to procure and sustain AI tools, as well as foundational AI education to equip frontline staff. For best practices in implementation, participants highlighted the need for standardized processes to evaluate AI tools, success measures to assess their impact, and guidance for communicating with patients about AI use. Participants emphasized that successful AI adoption goes beyond implementation, highlighting the importance of building internal structures to oversee AI use. They expressed the need to establish and operationalize AI governance policies within their organizations.

Throughout the program, a variety of strategies were developed to address barriers. For the most common barriers, all associated strategies were rated by participants as effective or minimally effective. Notably, the most effective strategies originated from the Hubs rather than individual Spokes, highlighting the value of centralized support in addressing implementation challenges.

### Contributions

The barriers identified in this study differ from those described in existing literature on technology adoption in SNOs, which has largely focused on broad, system-level challenges (11,13,15). The current findings identify granular, organization-specific barriers that reflect the realities of on-the-ground AI implementation. Furthermore, this research goes beyond identifying barriers by documenting associated mitigation strategies and reporting which strategies were effective. These insights can help SNO leaders anticipate potential challenges in AI adoption and proactively implement mitigation strategies, increasing the likelihood of successful AI adoption.

In addition, the current study provides preliminary insights into how AI implementation experiences in SNOs compare with those in larger, better-resourced HDOs, such as AMCs. Comparing the current findings with existing literature on AI implementation in AMCs reveals both common implementation challenges and context-specific differences (6,8,9,45). Several similarities emerged across settings. First, workforce training and education in using AI tools is critical, with SNOs placing particular emphasis on foundational AI knowledge for frontline staff that is ideally customized to their role (i.e., operational vs. clinical vs. technical). Second, effective communication regarding the use of AI tools is important for stakeholders, including end users and patients. Third, early engagement with end users and workflow optimization are essential to facilitate integration and reduce resistance to AI adoption. Finally, both settings recognize the importance of establishing standards and best practices for evaluating model performance and defining metrics for successful implementation.

Despite these commonalities, important differences were observed as well. Financial constraints, for example, were rarely reported in AMCs, whereas they represent a prominent barrier for SNOs. Relatedly, SNOs often lack negotiation power in establishing relationships with AI vendors. They face challenges in negotiating pricing, obtaining necessary information to assess vendors, identifying and obtaining expertise to support contracting, and securing adequate training materials for staff. Challenges with sustaining and scaling AI use are more pronounced in SNOs, in part because they typically rely on external vendors rather than developing tools in-house, creating a need for transparency around AI specifications and performance. Business Associate Agreements (BAAs) represent an important mechanism for holding vendors accountable to disclosed specifications and performance commitments, particularly regarding secondary and tertiary data use. Additionally, while AI governance structures are often already established at AMCs, SNOs face gaps in governance policies and processes, highlighting the need for capacity building in this area.

Taken together, these findings suggest that while foundational principles of AI implementation–such as evaluation, training, communication, and workflow integration–apply across settings, SNOs face distinct resource and structural challenges that require tailored strategies. By suggesting both similarities and differences, the current study advances understanding of context-specific AI implementation and informs practical approaches to supporting equitable adoption in resource-constrained organizations.

### Practical Implications

The current findings offer several practical lessons for promoting AI adoption in the safety net. First, the distributed Hub-and-Spoke model proved effective, as participation in the Practice Network Program increased AI readiness across all Spokes. Coordinating various types of expertise through the network of Hubs allowed Spokes to access guidance and support that would have been difficult to achieve individually. With this support, participating organizations all overcame barriers related to communicating with patients about AI tools and establishing AI governance. This finding demonstrates the value of diffusing knowledge and technical expertise from centralized experts to individual organizations. Peer learning–a component of the program–also emerged as an effective strategy. Facilitating safe spaces for SNOs to share common barriers, exchange knowledge, and develop mitigation strategies helped organizations navigate challenges collectively.

Despite these successes, three of the five most common barriers persisted at the conclusion of the program, underscoring that AI adoption in SNOs remains complex even with dedicated technical assistance. They included establishing systematic ways to evaluate the performance and impact of AI tools, equipping frontline staff with foundational AI knowledge and skills, and addressing financial constraints. Notably, some of these challenges–particularly evaluation and workforce training–have also been reported in larger, well-resourced HDOs (6,8,9,45). Addressing these challenges will require continued collaborative learning and sustained partnerships through active communities of practice, as demonstrated by HAIP’s model of convening multidisciplinary experts and SNOs. Professional societies will also play a critical role ensuring that training and continuing education programs build key capabilities among the workforce. For example, the American Medical Association offers a continuing medical education (CME) module focused on the ethical use of AI in clinical practice (47). Similarly, the Society of Teachers of Family Medicine provides a CME for medical students, primary care residents, faculty, and practicing primary care physicians to equip them with the competencies needed to integrate AI into clinical practice and promote the responsible and ethical use of these technologies (48). Such partnerships can support the co-development of shared resources–such as practical AI education materials, standardized approaches to AI evaluation, and systematic methods for assessing ROI–that address critical gaps and needs and advance safe, effective, and ethical AI implementation. Expanding these efforts to develop materials tailored to the full range of roles involved in AI implementation—including administrative leaders, operations staff, and data teams—would further strengthen organizational capacity across SNOs.

Finally, deliberate investment in foundational AI capacity is necessary to enable SNOs to overcome barriers, build partnerships, and support sustainable and scalable AI adoption. Even in well-resourced HDOs, AI adoption requires significant infrastructure and expertise. Targeted investment in SNOs can support safer, more effective, and ethically sound AI implementation. These findings highlight the need for action from state and federal actors to ensure equitable AI adoption in under-resourced health settings.

### Limitations and Future Directions

This study included a small sample of five SNOs. While limited in size, these organizations were representative of different regions across the U.S., and participants from each SNO brought diverse expertise. In addition, participants implemented diverse AI use cases, enabling the capture of distinct barriers associated with different types of AI tools. This approach provided rich, contextual, and in-depth insights into AI adoption in safety net settings.

The current findings underscore the value of technical assistance programs, such as the Practice Network Program, that create partnerships among multiple institutions to enable sustainable and scalable AI implementation. Building on these insights, future work by HAIP will focus on developing regional Practice Network Programs tailored to specific states, recognizing that differing policies and regulations shape AI adoption across regions. In this next phase, regional programs could adopt a “train-the-intermediary” model by recruiting and partnering with entities that already provide technical assistance to SNOs within a given region, such as HCCNs. Rather than HAIP supporting SNOs as in the current study, HAIP would provide targeted technical assistance and AI capacity-building support to HCCNs, equipping them with the knowledge, tools, and governance frameworks needed to guide AI adoption across their member organizations. Because HCCNs maintain close, ongoing relationships with SNOs and possess a deep understanding of their operational constraints, workforce capacity, patient populations, and regional regulatory environments, they are well positioned to translate AI guidance into contextually appropriate and actionable support. By strengthening the capabilities of these regional intermediaries, the programs can promote a more sustainable and scalable diffusion of AI expertise. Such programs can extend the lessons learned from this study and further support SNOs in navigating the complex landscape of AI in healthcare.

## Conclusion

This study provides an in-depth examination of real-world AI adoption in SNOs, identifying both barriers and effective mitigation strategies across the AI lifecycle. By centering the perspectives of individuals leading AI adoption within SNOs, the research amplifies voices that are often underrepresented in the health AI ecosystem, highlighting practical challenges around capacity, resources, and ethical guidance. The findings demonstrate that targeted technical assistance, particularly through a Hub-and-Spoke model, is critical for supporting AI adoption in resource-constrained settings. Peer learning, centralized expertise, and structured guidance enable organizations to navigate complex barriers more effectively than attempting adoption in isolation. At the same time, persistent challenges—such as local validation and post-deployment monitoring of AI tools, foundational AI education for frontline professionals, and financial constraints—underscore the need for ongoing collaborative support and deliberate investment in capacity-building. By providing granular, actionable insights, this study contributes to both practice and scholarship. Health system leaders can proactively anticipate barriers and implement mitigation strategies, while the research advances the emerging literature on AI technical assistance and implementation in under-resourced health settings. Looking forward, sustained investment, collaborative learning, and inclusion of SNO perspectives are essential to ensure that AI adoption supports safe, effective, and ethical care for the populations that rely on these critical SNOs.

## Data Availability

The authors have no additional data to report.

## Acknowledgement

We thank HAIP Corps Network members and Leadership Council who participated in the Practice Network Program as Hubs: Yasir Tarabichi, Corinne Stroum, David Vidal, Mark Lifson, Albert Karam, Steve Miff, William Ratliff, Hossein Soleimani, Autumn Zhu, Sara Murray, Vincent X. Liu, Elkin Salinas, Sofi Bergkvist, Jessica Ortiz, Alexandra Valladares, Zev Eigen, Sam Tyner-Monroe, and Elliot First. We are grateful for their time, contributions, and insights throughout the program. We also thank the Gordon and Betty Moore Foundation for funding the program.

